## Supplementary Tables for "Effectiveness of a multifaceted prevention programme for melioidosis in diabetics (PREMEL): a stepped-wedge cluster-randomised controlled trial"

**Supplementary Table 1: ICD-10-TM* codes and criteria used to determine that hospitals admissions were possibly related to infectious diseases.**

| ICD-10-TM | Diseases | n ** |
| --- | --- | --- |
| A00-B99 | Certain infectious and parasitic diseases*** | 1213 (36%) |
| D73.3 | Abscess of spleen | 20 (0.6%) |
| H10, H16, H44.0 | Eye infection | 14 (0.4%) |
| H60, H65, H66 | Infective otitis externa | 17 (1%) |
| I01, I33, I38, I39 | Endocarditis | 9 (0.3%) |
| J00-J06 | Acute upper respiratory infections | 88 (3%) |
| J09-J18 | Influenza and pneumonia**** | 372 (11%) |
| J20-J22 | Other acute lower respiratory infections | 51 (2%) |
| J85-J86 | Suppurative and necrotic conditions of the lower respiratory tract | 11 (0.3%) |
| K75.0 | Abscess of liver | 11 (0.3%) |
| K80, K81 | (Acute) cholecystitis | 104 (3%) |
| L00-L08, L30.3, L66.3, L66.4 | Infections of the skin and subcutaneous tissue***** | 452 (14%) |
| M00-M02, M86 | Bone and join infections | 91 (3%) |
| N13.6, N15.1,  N30, N39.0 | Urinary tract infection | 287 (9%) |
| R50.9 | Fever, unspecified | 241 (7%) |
| R57.2 | Septic shock | 143 (4%) |
| T79.3 | Post-traumatic wound infection, not elsewhere classified | 302 (9%) |
| T85.7 | Infection and inflammatory reaction due to other internal prosthetic devices, implants and grafts | 50 (1%) |
| Others | Having terms “fever, febrile, infect, abscess, pus, diarrhea, sepsis, pneumonia, cellulitis or diabetic foot” in a primary diagnosis | 1882 (56%) |
| Any ICD-10 codes | Culture positive for *Burkholderia pseudomallei* from any clinical specimens | 57 (2%) |
| Total | - | 3,335 |

* ICD-10-TM = International Statistical Classification of Diseases and Related Health Problems, 10th Revision, Thailand Modification ** Data are n (%) of 3,335 admissions involving infectious diseases. Codes were de-duplicated per hospital admission. *** Included A09 (diarrhoea and gastroenteritis of presumed infectious origin; n=582), A41 (sepsis, n=179), A24 (melioidosis, n=76), A05.9 (Bacterial foodborne intoxication, n=61), A49.9 (Bacterial infection, unspecified, n=42), etc. **** Included J18 (pneumonia, unspecified organism, n=264), J15 (bacterial pneumonia, not elsewhere classified, n=103), J209 (acute bronchitis, n=40), etc. ***** Included L03 (Cellulitis, n=251), L024 (cutaneous abscess, n=76) and L089 (Local infection of the skin and subcutaneous tissue, unspecified, n=52), etc.

**Supplementary Table 2: ICD-10-TM* codes and criteria used to determine that deaths were possibly related to infectious diseases.**

| ICD-10-TM | Diseases | n ** |
| --- | --- | --- |
| A00-B99 | Certain infectious and parasitic diseases*** | 108 (49.8%) |
| D73.3 | Abscess of spleen | 1 (0.5%) |
| H10, H16, H44.0 | Eye infection | 0 (0%) |
| H60, H65, H66 | Infective otitis externa | 0 (0%) |
| I01, I33, I38, I39 | Endocarditis | 3 (1.4%) |
| J00-J06 | Acute upper respiratory infections | 0 (0%) |
| J09-J18 | Influenza and pneumonia**** | 91 (41.9%) |
| J20-J22 | Other acute lower respiratory infections | 0 (0%) |
| J85-J86 | Suppurative and necrotic conditions of the lower respiratory tract | 4 (1.8%) |
| K75.0 | Abscess of liver | 1 (0.5%) |
| K80, K81 | (Acute) cholecystitis | 0 (0%) |
| L00-L08, L30.3, L66.3, L66.4 | Infections of the skin and subcutaneous tissue***** | 15 (7%) |
| M00-M02, M86 | Bone and join infections | 4 (2%) |
| N13.6, N15.1,  N30, N39.0 | Urinary tract infection | 29 (13%) |
| R50.9 | Fever, unspecified | 8 (4%) |
| R57.2 | Septic shock | 94 (43%) |
| T79.3 | Post-traumatic wound infection, not elsewhere classified | 8 (4%) |
| T85.7 | Infection and inflammatory reaction due to other internal prosthetic devices, implants and grafts | 13 (6%) |
| Others | Having terms “fever, febrile, infect, abscess, pus, diarrhea, sepsis, pneumonia, cellulitis or diabetic foot” in a primary diagnosis | 92 (42%) |
| Any ICD-10 codes | Culture positive for *Burkholderia pseudomallei* from any clinical specimens | 15 (7%) |
| Total | - | 217 |

* ICD-10-TM = International Statistical Classification of Diseases and Related Health Problems, 10th Revision, Thailand Modification ** Data are n (%) of 217 mortality involving infectious diseases. *** Included A41 (sepsis, n=53), A09 (diarrhoea and gastroenteritis of presumed infectious origin; n=18), A24 (melioidosis, n=14), etc. **** Included J18 (pneumonia, unspecified organism, n=55), J15 (bacterial pneumonia, not elsewhere classified, n=38), J90 (Pleural effusion, not elsewhere classified, n=10), etc. ***** Included L089 (Local infection of the skin and subcutaneous tissue, unspecified, n=13), etc.

**Supplementary Table 3. Factors associated with hospital admissions involving infectious diseases**

| **Factors** | **Adjusted rate ratio (95% CI)*** | **P value** |
| --- | --- | --- |
| Received the intervention per protocol | 0.90 (0.81-1.00) | 0.04 |
| Time period |  |  |
| Period 1 (Apr 2014 – Feb 2015) | 1.00 | <0.001 |
| Period 2 (Mar 2015 – Feb 2016) | 1.21 (1.05-1.39) |  |
| Period 3 (Mar 2016 – Feb 2017) | 1.51 (1.31-1.73) |  |
| Period 4 (Mar 2017 – Feb 2018) | 1.95 (1.69-2.25) |  |
| Period 5 (Mar 2018 – Dec 2018) | 2.13 (1.83-2.49) |  |
| Sex, female | 0.72 (0.64-0.80) | <0.001 |
| Age |  |  |
| 18 - <40 years | 1.00 | <0.001 |
| 40 - <55 years | 1.15 (1.02-1.31) |  |
| 55 – 65 years | 1.42 (1.24-1.63) |  |
| Diabetes duration |  |  |
| <5 years | 1.00 | <0.001 |
| 5 - <10 years | 1.22 (1.07-1.38) |  |
| ≥10 years | 1.99 (1.76-2.24) |  |
| HbA_1c_ level |  |  |
| <7.0 % | 1.00 | <0.001 |
| 7.0 - 8.0% | 1.06 (0.92-1.23) |  |
| >8.0 - 9.0% | 1.20 (1.02-1.41) |  |
| >9.0% | 2.27 (1.97-2.62) |  |

* CI=confidence interval. Estimated using a multivariable multilevel mixed-effect negative binomial regression model with a random effect for PCU (n=9,056 diabetic patients)

**Supplementary Table 4. Factors associated with mortality**

| **Factors** | **Adjusted rate ratio (95% CI)** | **P value** |
| --- | --- | --- |
| Received the intervention per protocol | 0.56 (0.44-0.71) | <0.001 |
| Time period |  |  |
| Period 1 (Apr 2014 – Feb 2015) | 1.00 | <0.001 |
| Period 2 (Mar 2015 – Feb 2016) | 1.39 (0.93-2.08) |  |
| Period 3 (Mar 2016 – Feb 2017) | 2.07 (1.40-3.07) |  |
| Period 4 (Mar 2017 – Feb 2018) | 3.01 (2.04-4.44) |  |
| Period 5 (Mar 2018 – Dec 2018) | 3.42 (2.27-5.15) |  |
| Sex, female | 0.42 (0.35-0.51) | <0.001 |
| Age |  |  |
| 18 - <40 years | 1.00 | <0.001 |
| 40 - <55 years | 1.40 (1.08-1.81) |  |
| 55 – 65 years | 2.09 (1.60-2.72) |  |
| Diabetes duration |  |  |
| <5 years | 1.00 | <0.001 |
| 5 - <10 years | 1.30 (1.02-1.66) |  |
| ≥10 years | 2.03 (1.62-2.56) |  |
| HbA_1c_ level |  |  |
| <7.0 % | 1.00 | <0.001 |
| 7.0 - 8.0% | 0.82 (0.62-1.08) |  |
| >8.0 - 9.0% | 1.16 (0.87-1.54) |  |
| >9.0% | 1.40 (1.09-1.80) |  |

* CI=confidence interval. Estimated using a multivariable multilevel mixed-effect Poisson regression model with a random effect for PCU (n=9,056 diabetic patients)

**Supplementary Table 5. Factors associated with overall melioidosis**

| **Factors** | **Adjusted rate ratio (95% CI)*** | **P value** |
| --- | --- | --- |
| Received the intervention per protocol | 0.74 (0.38-1.44) | 0.37 |
| Time period |  |  |
| Period 1 (Apr 2014 – Feb 2015) | 1.00 | 0.41 |
| Period 2 (Mar 2015 – Feb 2016) | 0.73 (0.34-1.54) |  |
| Period 3 (Mar 2016 – Feb 2017) | 1.34 (0.65-2.73) |  |
| Period 4 (Mar 2017 – Feb 2018) | 0.87 (0.38-2.08) |  |
| Period 5 (Mar 2018 – Dec 2018) | 0.95 (0.34-2.67) |  |
| Sex, female | 0.28 (0.18-0.44) | <0.001 |
| Age |  |  |
| 18 - <40 years | 1.00 | 0.12 |
| 40 - <55 years | 0.74 (0.44-1.22) |  |
| 55 – 65 years | 0.52 (0.28-0.98) |  |
| Diabetes duration |  |  |
| <5 years | 1.00 | 0.005 |
| 5 - <10 years | 1.21 (0.66-2.21) |  |
| ≥10 years | 2.37 (1.36-4.12) |  |
| HbA_1c_ level |  |  |
| <7.0 % | 1.00 | <0.001 |
| 7.0 - 8.0% | 1.71 (0.77-3.79) |  |
| >8.0 - 9.0% | 0.90 (0.33-2.45) |  |
| >9.0% | 3.42 (1.64-7.15) |  |

* CI=confidence interval. Estimated using a multivariable multilevel mixed-effect Poisson regression model with a random effect for PCU (n=9,056 diabetic patients)

**Supplementary Table 6. Potential cost of the prevention programme of melioidosis in diabetic patients in Ubon Ratchathani, Thailand**

| **Description** | **Cost** |
| --- | --- |
| **Material**  For each participant, a color calendar together with an individual photograph acting as a reminder was provided. | **1$ per diabetic patient** |
| **Compensation**  For participants to offset the time and inconvenience of participation, and as incentive to participate | **2$ per diabetic patient**  The study provided a pair of long socks and a bottle of baby powder and a 2-litre plastic ice bucket. |
| **Human resource**  Based on our experience, each session requires 2 research assistants. Each session takes about 1 hour. However, we assume that each session requires 3 hours of each research assistant to conduct in order to compensate for preparation and training. Therefore, each session requires a total of 6 working hours of a research assistant. We also estimated that each research assistant salary was about 500$/month (minimum salary recommended by the government of Thailand for new employees with a bachelor's degree was 15,000 THB/month; and the 1$=30THB), and each research assistant work 160 hours per month. Therefore, the cost of one working hour of a research assistant is about 3.125$ | **2$ per diabetic patient**  Using the assumptions, we estimated that the labor cost would be about 19$ per session (3.125$ per working hour per research assistant * 6 working hours of research assistant). If there are on average 10 diabetic patient per session, the labor cost could be about 2$ per diabetic patient. |
| **Total** | **5$ per diabetic patient** |
